# Evaluation of Closed and Open Large Language Models in Pediatric Cardiology Board Exam Performance

**DOI:** 10.1101/2025.06.28.25330485

**Authors:** Nino Nikolovski, Conall T. Morgan, Michael N. Gritti

## Abstract

**Introduction:** Large language models (LLMs) have gained traction in medicine, but there is limited research comparing closed- and open-source models in subspecialty contexts. This study evaluated ChatGPT-4.0o and DeepSeek–R1 on a pediatric cardiology board-style examination to quantify their accuracy and discuss clinical and educational utility.

**Methods:** ChatGPT-4.0o and DeepSeek–R1 were used to answer 88 text-based multiple-choice questions across 11 pediatric cardiology subtopics from a *Pediatric Cardiology Board Review* textbook. DeepSeek–R1’s processing time per question was measured. Statistical analyses for model comparison were conducted using an unpaired two-tailed t-test, and bivariate correlations were assessed using Pearson’s r.

**Results:** ChatGPT-4.0o and DeepSeek–R1 achieved 70% (62/88) and 68% (60/88) accuracy, respectively (p=0.79). Subtopic accuracy was equal in 5 of 11 chapters, with each model outperforming its counterpart in 3 of 11. DeepSeek–R1’s processing time negatively correlated with accuracy (r = –0.68, p = 0.02).

**Conclusion:** ChatGPT-4.0o and DeepSeek–R1 approached the passing threshold on a pediatric cardiology board examination, with comparable accuracy and potential for open-source models to enhance clinical and educational outcomes while supporting sustainable AI development.

## Introduction

The integration of artificial intelligence (AI) into medicine for enhancing clinical workflows and quality of care has expedited with the emergence of large language models (LLMs).^1,2^ LLMs are machine learning models capable of performing complex reasoning tasks and generating contextually accurate responses.^2^ Proprietary, closed-source models such as OpenAI’s Chat Generative Pre-Trained Transformer (GPT) have gained traction in medicine, partly due to their ability to demonstrate accuracy and feasibility in radiologic decision making,^2,3^ and provide higher quality and more empathetic responses compared to physicians.^4^ The most recent iteration, ChatGPT-4.0 omni (o), has exhibited substantial improvements in accuracy compared to its predecessors.^5^ ChatGPT-3.5 and ChatGPT-4.0 were recently compared using a pediatric cardiology educational knowledge assessment and despite both versions failing to achieve a passing score, it was identified that the successor had superior performance overall and within respective subtopics.

The architecture and training datasets of closed-source models are obscured from public knowledge due to commercial competition, preservation of intellectual property, and protection from adversarial attacks.^6^ Proprietary LLMs have been criticized for the lack of accessibility and financial resources required for training the model and commercial use.^1^ Moreover, their application in healthcare contexts may be limited by relying on a particular version of the model.^2^ Conversely, open-source, free to access alternatives like DeepSeek’s DeepSeek–R1 have emerged, aiming to match proprietary LLM performance with transparency, accessibility, and resource efficiency enabling users to visualize and customize the model’s source code.^1,2^ While open-source LLMs have received less attention in the medical literature, Temsah *et al*. highlight the capacity for continuous learning and discipline-specific modulation of these models through integration with publicly available datasets. This facilitates alignment of the model’s training with novel scientific research and current medical curricula, and by extension great potential for adapting to the demands of an everchanging medical landscape.

While LLMs may have a transformative role for patients and physicians, they must achieve a level of accuracy that can be relied upon consistently for clinical implementation without inherent biases or hallucinations.^1^ There are limited comparative evaluations of proprietary and open-source LLMs in subspecialty medical disciplines such as pediatric cardiology. To address this knowledge gap and facilitate the clinical integration of AI, the present study evaluated the performance of ChatGPT-4.0o and DeepSeek–R1 on a multiple-choice examination modeled after a pediatric cardiology board examination. The objectives of the present study are: (1) to compare the overall and topic-specific accuracy between models, and (2) to determine whether closed and/or open-sourced LLMs are capable of meeting the 70% passing standard for board certification in pediatric cardiology to indicate potential readiness for educational and clinical use.

## Methods

Reproducing the methodology from Gritti *et al*., we used a dataset of 88 multiple choice questions taken verbatim from *Pediatric Cardiology Board Review*^7^ by Eidem *et al*. ChatGPT-4.0o and DeepSeek–R1 were used to answer questions obtained from the question bank. The default mode for ChatGPT-4.0o was used with ChatGPT plus, and this chatbot was trained on available data up to October 2023. DeepSeek–R1’s “DeepThink (R1)” feature was toggled on for every question input, enabling the user to observe the open-source model’s thought process based on its training and quantify processing time per question. We arbitrarily used the first eight “text-only” questions from each of the 11 eligible chapters in the textbook for a total of 88 questions per test. We excluded questions that had associated images and other modalities such as sound.

Each chapter contained eight questions regarding one of the following 11 pediatric cardiology topics: *Cardiac Anatomy and Physiology, Congenital Cardiac Malformations, Diagnosis of Congenital Heart Disease, Cardiac Catheterization and Angiography, Non-invasive Cardiac Imaging, Electrophysiology Questions for Paediatrics, Exercise Physiology and Testing, Outpatient Cardiology, Cardiac Intensive Care and Heart Failure, Cardiac Pharmacology* and *Surgical Palliation and Repair of Congenital Heart Disease*. Questions from other chapters were excluded if they were not specific to pediatric cardiology (i.e., statistics).

Each question was entered as a separate new prompt, and previous conversations were cleared to ensure no previous information affected the chatbot’s answers. Each question was entered into ChatGPT-4.0o and DeepSeek–R1 with identical wording. Each answer was then manually reviewed by members of our team (N.N, C.T.M, and M.N.G) to ensure that the chatbots had answered the question. If either LLM deemed that none, multiple or all the answers were correct, when this was not one of the multiple choice options, it was scored as incorrect. The answer key provided by the textbook was used to verify correct answers and both ChatGPT-4.0o and DeepSeek–R1 were given a score out of 8 for each question they correctly answered in each chapter. Normality of the data was confirmed with a Shapiro-Wilk test. Statistical analyses were conducted on SPSS 20 (IBM Software) using an unpaired two-tailed t-test and Pearson’s r correlation, with a p-value less than 0.05 being statistically significant. Due to the nature of this study, no research ethics approval from our institution was obtained.

## Results

### Overall Model Performance

ChatGPT-4.0o and DeepSeek–R1 were used to answer 88 text-based multiple choice questions from the *Pediatric Cardiology Board Review*^7^ textbook. Data are reported as a mean percentage (%) of total questions answered correctly. ChatGPT-4.0o answered 62 questions, or 70% correctly, while DeepSeek–R1 answered 60 questions, or 68% correctly. While ChatGPT-4.0o answered 2 more questions correctly relative to DeepSeek–R1, model performance was not statistically different (Figure 2; p = 0.79).

### Subtopic Stratification

#### ChatGPT 4.0o

ChatGPT-4.0o data were also stratified by subtopic according to 11 distinct chapters and are presented as number of questions answered correctly out of 8, and mean±SD. With regard to stratified subtopic data, ChatGPT-4.0o answered 5.6±1.7 questions correctly. ChatGPT-4.0o performed best on *Chapter 8: Exercise Physiology & Testing* (8/8 questions answered correctly, 100%) and worst on *Chapters 3 and 4: Diagnosis of Congenital Heart Disease and Cardiac Catheterization & Angiography*, respectively (Table 1; 3/8 questions answered correctly in both, 38%). ChatGPT-4.0o answered 70±21% questions correctly.

**Table 1:** Number of correctly answered questions (out of 88) by ChatGPT-4.0o and DeepSeek– R1 in each of the 11 chapters of the *Pediatric Cardiology Board Review* textbook.^7^.

|  | ChatGPT-4.0o | DeepSeek-R1 |
| --- | --- | --- |
| *Chapter 1: Cardiac Anatomy and Physiology (/8)* | **4** | **5** |
| *Chapter 2: Congenital Cardiac Malformations (/8)* | **6** | **6** |
| *Chapter 3: Diagnosis of Congenital Heart Disease (/8)* | **3** | **3** |
| *Chapter 4: Cardiac Catheterization and Angiography (/8)* | **3** | **3** |
| *Chapter 5: Non-invasive Cardiac Imaging (/8)* | **7** | **7** |
| *Chapter 6: Electrophysiology Questions for Paediatrics (/8)* | **7** | **7** |
| *Chapter 7: Exercise Physiology and Testing (/8)* | **8** | **5** |
| *Chapter 8: Outpatient Cardiology (/8)* | **7** | **6** |
| *Chapter 9: Cardiac Intensive Care and Heart Failure (/8)* | **6** | **5** |
| *Chapter 10: Cardiac Pharmacology (/8)* | **5** | **6** |
| *Chapter 11: Surgical Palliation and Repair of Congenital Heart Disease (/8)* | **6** | **7** |
| *Total (/88)* | **62** | **60** |
| *Score on entire examination* | **70%** | **68%** |

#### DeepSeek–R1

DeepSeek–R1 data were also stratified by subtopic according to 11 distinct chapters and are presented as number of questions answered correctly out of 8, and mean±SD. With regard to stratified subtopic data, DeepSeek–R1 answered 5.5±1.4 questions correctly. DeepSeek–R1 performed best on *Chapters 5, 6, and 11: Non-invasive Cardiac Imaging, Electrophysiology, and Surgical Palliation & Repair of Congenital Heart Disease*, respectively (Table 1; 7/8 questions answered correctly in all, 88%). DeepSeek–R1 answered 68±18% questions correctly. Using DeepSeek–R1’s DeepThink (R1) feature, this model outlined its thought process for 51±15 s per subtopic, with a range of 12–218 s for individual questions. The model thought the slowest on average for 67.6 s in *Chapter 3: Diagnosis of Congenital Heart Disease*, and the fastest on average for 36.0 s in *Chapter 1: Cardiac Anatomy & Physiology*. Pearson’s correlation analysis revealed a strong negative correlation (Figure 1; r = –0.68, p = 0.02) between number of questions answered correctly per chapter and length of DeepThink (R1) processing time.

**Figure 1:**
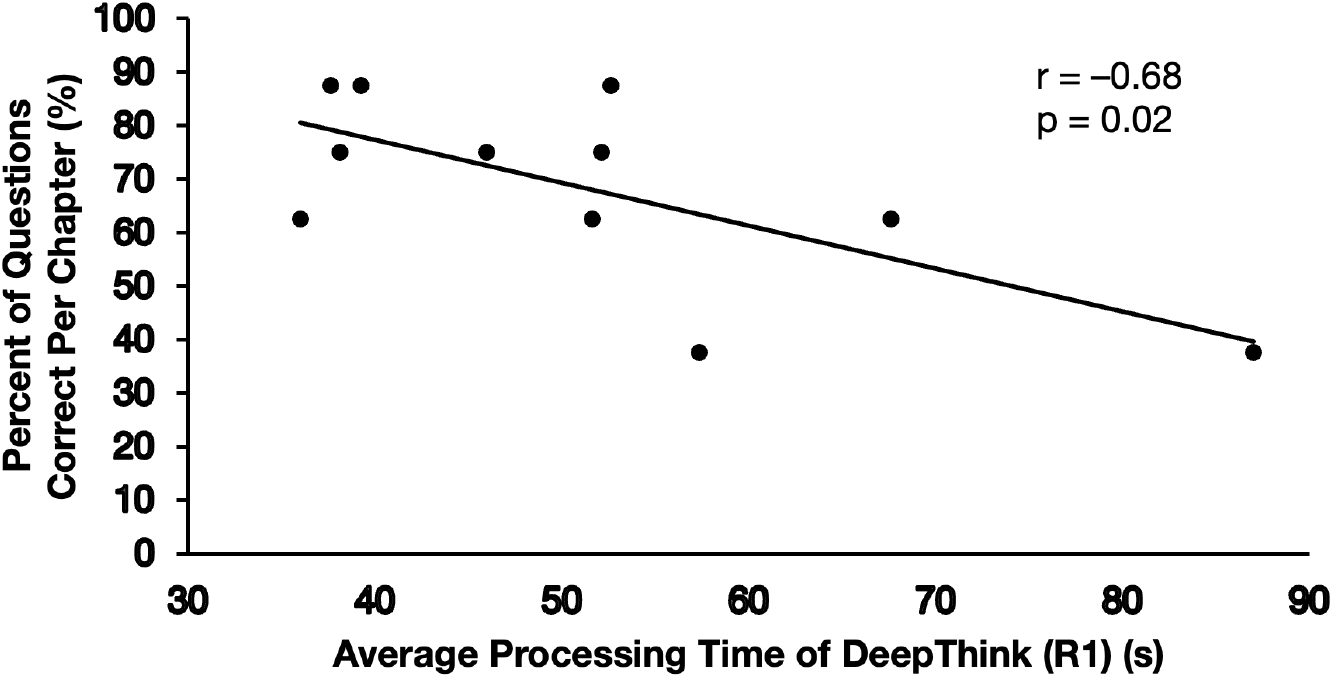
Relationship between average processing time (seconds) and percentage (%) of questions answered correctly by DeepSeek–R1 across 11 chapters in the *Pediatric Cardiology Board Review* book.^7^ Data displays a statistically significant strong negative correlation (r = –0.68, p = 0.02).

**Figure 2:**
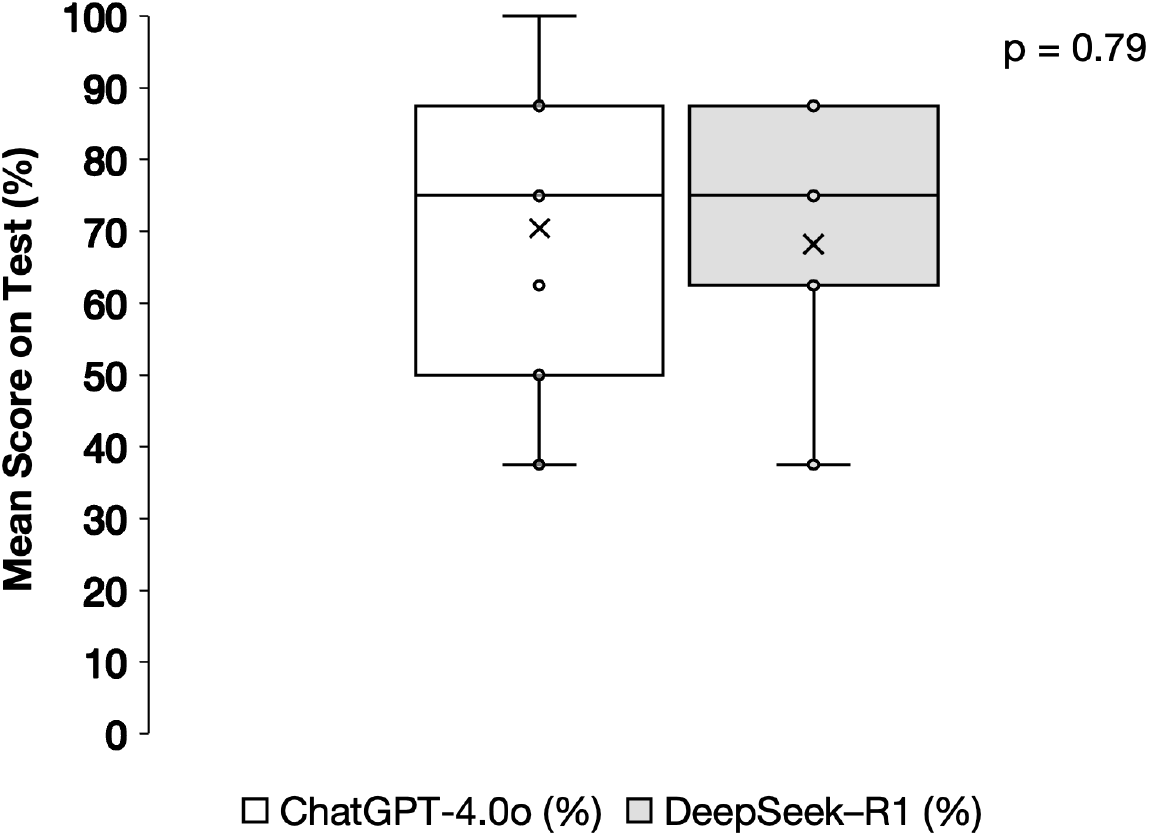
ChatGPT-4.0o and DeepSeek–R1 were each asked the same eight text based multiple choice questions from each of 11 chapters (n=88 for both) in the *Pediatric Cardiology Board Review*.^7^ “X” indicates mean scores. Data were analyzed with an unpaired t-test with no statistically significant difference between models (p = 0.79).

### ChatGPT-4.0o vs. DeepSeek–R1

ChatGPT-4.0o and DeepSeek–R1 performed equally on 5 out of 11 chapters. Specifically, *Chapter 2: Congenital Cardiac Malformations* (6/8 correct, 75%), *Chapter 3: Diagnosis of Congenital Heart Disease* (3/8 correct, 38%), *Chapter 4: Cardiac Catheterization & Angiography* (3/8 correct, 38%), *Chapter 5: Non-invasive Cardiac Imaging* (7/8 correct, 88%), and *Chapter 6: Electrophysiology* (7/8 correct, 88%). ChatGPT-4.0o outperformed DeepSeek–R1 on 3 out of 11 chapters. Specifically, *Chapter 7: Exercise Physiology & Testing* (8/8 correct, 100% vs. 5/8 correct, 63%), *Chapter 8: Outpatient Cardiology* (7/8 correct, 88% vs. 6/8 correct, 75%), *Chapter 9: Cardiac Intensive Care & Heart Failure* (6/8 correct, 75% vs. 5/8 correct, 63%). Conversely, DeepSeek–R1 outperformed ChatGPT-4.0o in 3 chapters. Specifically, *Chapter 1: Cardiac Anatomy & Physiology* (5/8 correct, 63% vs. 4/8 correct, 50%), *Chapter 10: Cardiac Pharmacology* (6/8 correct, 75% vs. 5/8 correct, 63%), and *Chapter 11: Surgical Palliation & Repair of Congenital Heart Disease* (7/8 correct, 88% vs. 6/8 correct, 75%). These results are summarized in Table 1.

## Discussion

This study aimed to compare and evaluate the performance of a closed-source LLM, ChatGPT-4.0o, to an open-source LLM, DeepSeek–R1, on a pediatric cardiology board-style examination to assess their potential utility in medical education and clinical integration. The central findings of the present study are: 1) ChatGPT-4.0o achieved a higher overall accuracy (70%) than DeepSeek–R1 (68%) with no statistically significant difference between the models, and 2) Both models performed at or near the accepted passing threshold of 70%. These findings offer novel evidence that proprietary and open-source LLMs can meet competency thresholds in a specialized medical discipline; however, further validation and reliability assessments are pre-requisites for their implementation in clinical and medical education contexts.

The marginally superior, albeit statistically insignificant (p=0.79), performance of ChatGPT-4.0o relative to DeepSeek–R1 has practically significant implications. Achieving the designation of board-certified pediatric cardiologist is binary based on the multiple choice examination as one either passes with 70%+ or is unsuccessful below this threshold. The ability of ChatGPT-4.0o to achieve a passing grade is in alignment with evidence that closed-source LLMs traditionally demonstrate superior performance on AI benchmarking protocols used for comparative analyses between different model architectures.^2,6^ These performance differences in dimensions such as linguistic proficiency and domain-specific adaptability can be ascribed to closed-sourced LLMs’ training on extensive proprietary datasets. This elaborate pre-training enables proficiency in broader generalizability and multi-step complex reasoning required in medical subspecialties such as pediatric cardiology. However, the present findings also align with data revealing that open-sourced LLMs are rapidly bridging the performance divide through improvements in domain-specific fine-tuning.^2^ This bridging is evidenced through DeepSeek–R1 performing equally on 5/11 and better on 3/11 subtopics in a pediatric cardiology knowledge assessment. The inter-subtopic variability where each model excelled in different domains suggests that the underlying training data differ and provides an avenue for future investigations.

A strength of the present study is the measurement of processing time through DeepSeek– R1’s “DeepThink” feature to gain preliminary insight into the model’s reasoning capacity across pediatric cardiology subtopics. DeepSeek–R1 exhibited a strong negative correlation between processing time and accuracy per subtopic (Figure 1). This finding suggests that increased contemplation was associated with decreased performance. This inverse relationship provides proof-of-concept that LLMs have difficulty with multi-step reasoning problems which necessitate more deliberation compared to factual recall questions. While ChatGPT-4.0o does not enable insight into its thought process unless directly prompted, processing time in open-sourced LLMs may serve as a valuable diagnostic tool for identifying model deficiencies. These inadequacies may be corrected through domain-specific customization. For instance, Savage *et al*. emphasize the need for radiology-specific evaluation benchmarks to establish evidence-based standards for clinical use–a suggestion that can be applied to pediatric cardiology to augment quality of care.

Open-source LLMs are advantageous for their customizability and comparatively smaller size. Despite their substantiated utility in clinical applications, LLMs heighten the climate crisis through carbon emissions.^8^ It is imperative to seek solutions that enable us to maximize the value of AI while minimizing environmental impact. The smaller size of open-source LLMs attenuates the environmental footprint associated with the energy-intensive requirements of closed-source counterparts. LLM size is based on the number of parameters it is trained on, analogous to neurons and their synaptic connections enabling us to think and make decisions. While more parameters signify improved performance, the trade-off is a proportionally higher energy consumption. Doo *et al*. determined that smaller LLMs trained specifically for radiograph interpretation have greater diagnostic accuracy while consuming seven times less energy than general-purpose models. However, the improved accuracy of medically specialized models relative to general-purpose models may be contentious.^3^ Collectively, these findings suggest that domain-specific models should be developed in medical subspecialties to balance technological advancement and environmental sustainability without relying on subscription-based models. As pediatric cardiology demands multimodal image interpretation for effective diagnosis and treatment,^9,10^ open-source fine-tuned LLMs have great potential to be integrated into postgraduate training to provide trainees with the tools to be successful physicians.^3^

## Conclusion

In conclusion, ChatGPT-4.0o and DeepSeek–R1 performed at or near the 70% passing threshold on a pediatric cardiology assessment. While DeepSeek–R1 marginally missed this threshold, the statistical indifference in accuracy suggests that open-source models are narrowing performance disparities in medical subspecialty contexts. Processing time may serve as a diagnostic metric for LLM performance and gaps. Future research in open-source AI should direct attention to developing models specific to the multimodal demands of pediatric cardiology for advancing quality of care and medical education while minimizing environmental impact.

## Data Availability

All data produced in the present study are available upon reasonable request to the authors

## Acknowledgements

Not applicable.

## Statements and Declarations

### Ethical considerations

Not applicable.

### Consent to participate

Not applicable.

### Consent for publication

Not applicable.

### Data availability

Not applicable.

